# Cadmium and Lead Attributable Burden of Cancers in Skipjack (*Katsuwonus pelamis*) and Yellowfin (*Thunnus albacares*) in Ghana

**DOI:** 10.1101/2024.12.19.24319399

**Authors:** J. A. Nkansah, H.E. Lutterodt, A. Dodoo, I.W. Ofosu

## Abstract

Toxic heavy metal contamination in fish sources poses significant health risks, particularly in populations that rely on fish as a primary dietary component. Thus, this study aimed to assess the concentrations of cadmium (Cd) and lead (Pb) in selected tuna species and evaluate the associated health risks and cancer burdens to consumers in Ghana through a systematic review and meta-analysis of data covering the period 2015 to 2019. A Disability Adjusted Life Year (DALY) approach was used to quantify the health burden with secondary data from the Institute of Health Metrics and Evaluation (IHME). Median contaminant exposure was 1.26 × 10^-5^ mg/kg(bw)-day for Cd, and 1.04 × 10^-4^ mg/kg(bw)-day for Pb. The highest DALY was found in the elderly age group (55-89) and it ranged from 10^-5^ - 10^-3^. The results indicate that in the elderly age group, stomach cancer was the most prevalent for both males and females, reflecting a significant cancer burden. For elderly males, prostate cancer was also a severe concern, contributing to the overall cancer burden, although its prevalence was lower than that of stomach cancer. In elderly females, however, pancreatic cancer was observed to have a higher prevalence compared to males, indicating gender-specific differences in cancer risk among older adults. The findings underscore the need for public health interventions to mitigate the risks of heavy metal contamination in fish.

## 1.0 Introduction

Some heavy metals, such as cadmium and lead, can bioaccumulate in fish, especially those inhabiting polluted waters (1). This observation is a significant public health threat for the many communities which consume fish as their primary protein source. Some studies show that the deleterious effects of these pollutants disproportionately impact many impoverished communities worldwide (2). While the exact numbers concerning which poor communities eat fish and how many tons of fish are consumed can be debated, seafood is a staple or supplemental part of the diet for most impoverished populations. The intake of high quantities of fish which are contaminated with heavy metals can be a severe public health problem (3).

These deleterious pollutants in aquatic environments primarily originate from anthropogenic activities and natural sources such as the weathering of rocks and volcanic eruptions. However, human activities, including mining, can significantly accelerate the release of these contaminants, increasing their bioavailability (4). Industrial processes often release wastewater contaminated with heavy metals. Additionally, agricultural runoff is another major contributor because applying fertilizers and pesticides can lead to the leaching of heavy metals into nearby water bodies (5). Atmospheric deposition is also a major concern, particularly following the proliferation of heavy industrial activities. Subsequently, precipitation can transport these pollutants from the atmosphere to aquatic environments.

The bioaccumulation of heavy metals in aquatic ecosystems significantly threatens wildlife and human health. These contaminants can enter water bodies through various sources, including industrial waste, agricultural runoff, and atmospheric deposition. Once in the water, they can be absorbed by aquatic organisms, leading to bioaccumulation and biomagnification. This process can result in the concentration of heavy metals in higher trophic levels, ultimately affecting top predators and humans who consume contaminated seafood (6). Studies have shown that consuming contaminated seafood is the primary exposure route for cadmium and lead from fish to humans (7). When fish inhabit waters polluted with these heavy metals, they can absorb them through their gills and skin. Consequentially, these metals accumulate, particularly in the liver and kidneys. If humans consume such contaminated fish, toxic heavy metals can be transferred to their bodies, potentially leading to various health problems (8).

The adverse health effects of chronic exposure to heavy metals such as cadmium and lead include renal failure, neurotoxicity, cardiovascular disease (9) and reproductive abnormalities (10). Heavy metals such as cadmium, lead, and mercury can act as metalloestrogens, binding to estrogen receptors and disrupting normal hormonal functions, potentially contributing to the development of hormone-related diseases (11). Heavy metal exposure is also potentially linked to cancer development through oxidative stress, disruption of cell division, inhibition of apoptosis, and interference in DNA repairs (12).

Accurate human exposure assessment is required to effectively minimize the intake of toxic heavy metals and promote sustainable fish consumption. A study conducted in Poland found that while commercial fishery products generally contained safe levels of cadmium and lead, some products, such as salads and marinated fish, had higher concentrations of these metals (13). Despite this observation, the overall consumption of fish and fishery products in Poland was considered safe. Similarly, fish consumption rates across different demographics with higher consumption rates have been observed in coastal regions and among populations with traditional fish-based diets (14).

These findings highlight the importance of targeted public health interventions to address potential health risks associated with fish consumption in these populations.

Recent studies such as the Healthy Ageing and Biomarkers Cohort Study (HABCS) in China, have been conducted to investigate heavy metal bioaccumulation in human populations (15). These studies have revealed that older age groups are particularly vulnerable to higher levels of toxic metals like cadmium and lead. In Bosnia and Herzegovina, (16) tested for cadmium, mercury, and lead levels in seven commercial fish species and reported health risks for vulnerable populations. These underscore the need for continuous monitoring and targeted interventions to mitigate the health risks associated with toxic heavy metal exposure, particularly in ageing populations.

Like other developing countries, Ghana also faces a growing threat to public health due to the reported increasing levels of toxic heavy metals in aquatic ecosystems (17). Despite mounting evidence of the health risks associated with toxic heavy metal exposure, inadequate measures have been implemented to protect public health (18). The consequences of this gap extend beyond individual health to impact healthcare systems and hinder progress towards achieving the Sustainable Development Goals (SDGs). This present study aimed to investigate the occurrence of cadmium and lead in selected tuna species landed and consumed in Ghana and to determine the associated health risks and burden of cancers.

## 2.0 Methodology

### 2.1 Elements of exposure to cadmium and lead

The study examined the exposure of consumers in Ghana to cadmium and lead through their consumption of skipjack tuna (*Katsuwonus pelamis*) and yellowfin tuna (*Thunnus albacares*). Factors considered include the median concentrations of these toxic metals in the fish, a body weight of 60 kg (19), and the average quantity of fish consumed in Ghana, which has been reported to be 0.0685 kg/day (20).

#### 2.1.1 Cadmium and lead concentrations in fish

**Describing the historicity of cadmium and lead concentration data:** Cadmium and lead concentrations used in this study were archived historical data determined from samples of skipjack tuna and yellowfin tuna caught in the Food and Agriculture Organization (FAO) Major Fishing Area Zone 34 (21), illustrated in Figure 1.

**Figure 1:**
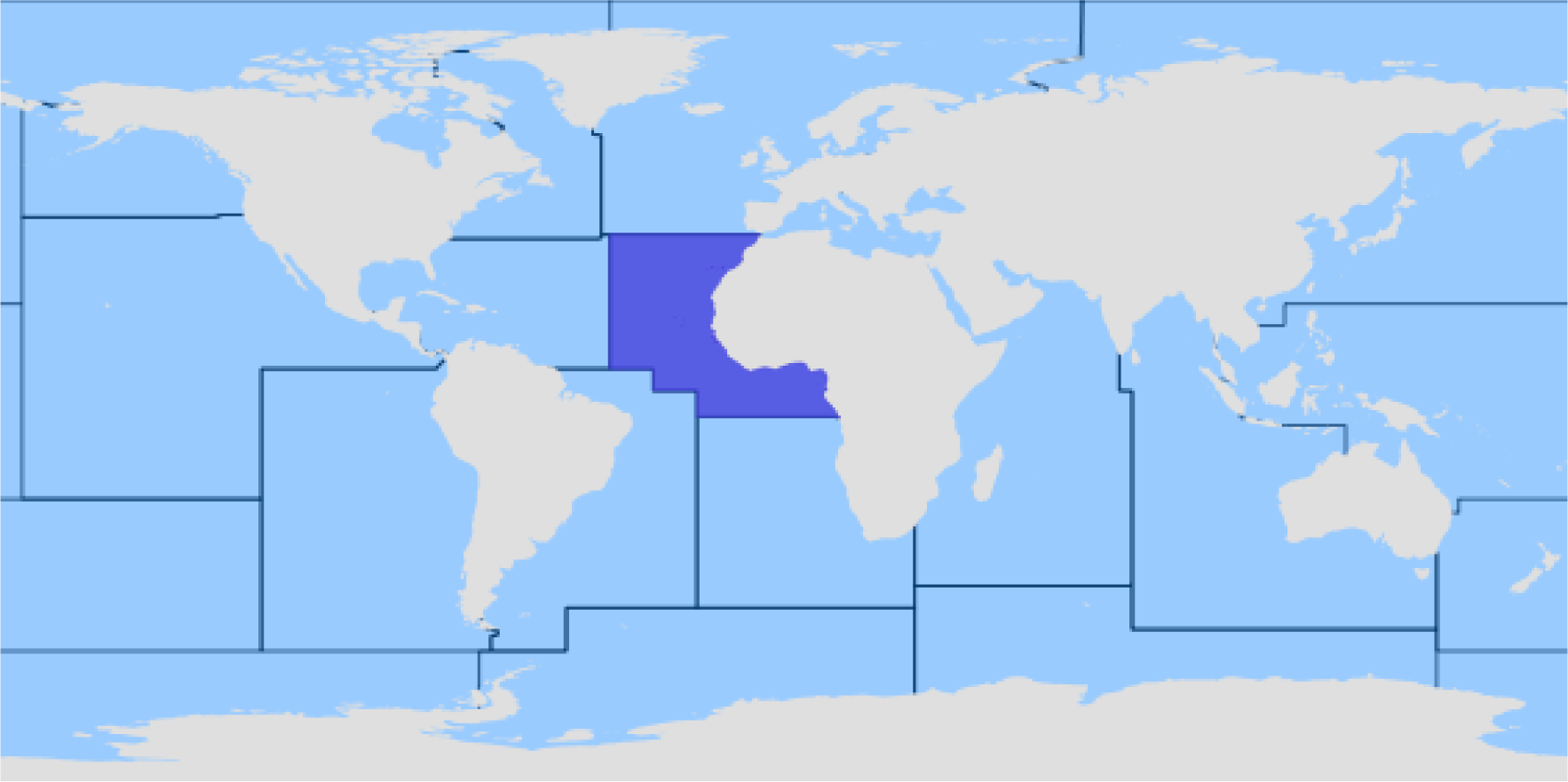
Boundaries of the subareas of the Atlantic showing a central fishing area (FAO Zone 34)

The fish samples were collected from 2015 to 2019 by trained inspectors from Ghana’s Competent Authority for Fish and Fishery Products, adhering to standardized procedures. The analyses were conducted in an accredited EN ISO/IEC 17025 laboratory. Cadmium and lead levels were analyzed using Inductively Coupled Plasma Mass Spectrometry (ICP-MS) calibrated with standard solutions and operated under specific conditions to ensure accurate results (22).

**Meta-analysis:** The five-year historical data collected presented potential challenges due to data inconsistency and the need for control groups. To address the issues, it was crucial to assess data quality by carefully considering the inclusion of studies involving control groups, interpreting effect sizes in context, and conducting sensitivity analyses. Data studies that follow these steps present meta-analyses that provide a more reliable and informative assessment of the relevant outcomes (23). The meta-analysis examined cadmium and lead concentrations in selected tuna species samples collected between 2015 and 2019. Heterogeneity was assessed using a chi-squared test, and subgroup analyses were conducted based on heavy metal type and year of data capture. The pooled proportions of cadmium and lead were estimated using the random effects model according to the DerSimonian Laird approach, and 95% confidence intervals were calculated (24). The analysis results were presented in a stratified forest plot, graphically depicting the estimated effect sizes, their corresponding confidence intervals for each study, and the overall pooled estimate. The forest plot also highlighted the heterogeneity among the studies and the potential for subgroup differences (25). These findings provided valuable insights into the trends and variability of cadmium and lead contamination in tuna samples over the study period.

### 2.2 Collection of health metrics data

Health metric data was obtained from the Institute for Health Metrics and Evaluation (26). The data collected focused on mortality, prevalence, years of life lost (YLL), and years lived with disability (YLD) for both males and females. The collection included five cancers attributed to cadmium and two cancers attributed to lead. The age groups analyzed were 15-19, 20-54, and 55-89. The data was collected for the years 2015 to 2019.

### 2.3 Data syntheses

Figure 2 presents a conceptual framework for assessing cadmium and lead health risks and cancer burdens in some selected fish species. It outlines the steps in quantifying the disease burden attributable to these hazards, illustrating the interconnectedness of critical factors such as their exposure, health metrics, consumer population, and cancer slope factors. The framework helps visualize and quantify the burden of these toxic heavy metals, providing a clearer understanding of the relationship between exposure, hazard, and health outcomes.

**Figure 2:**
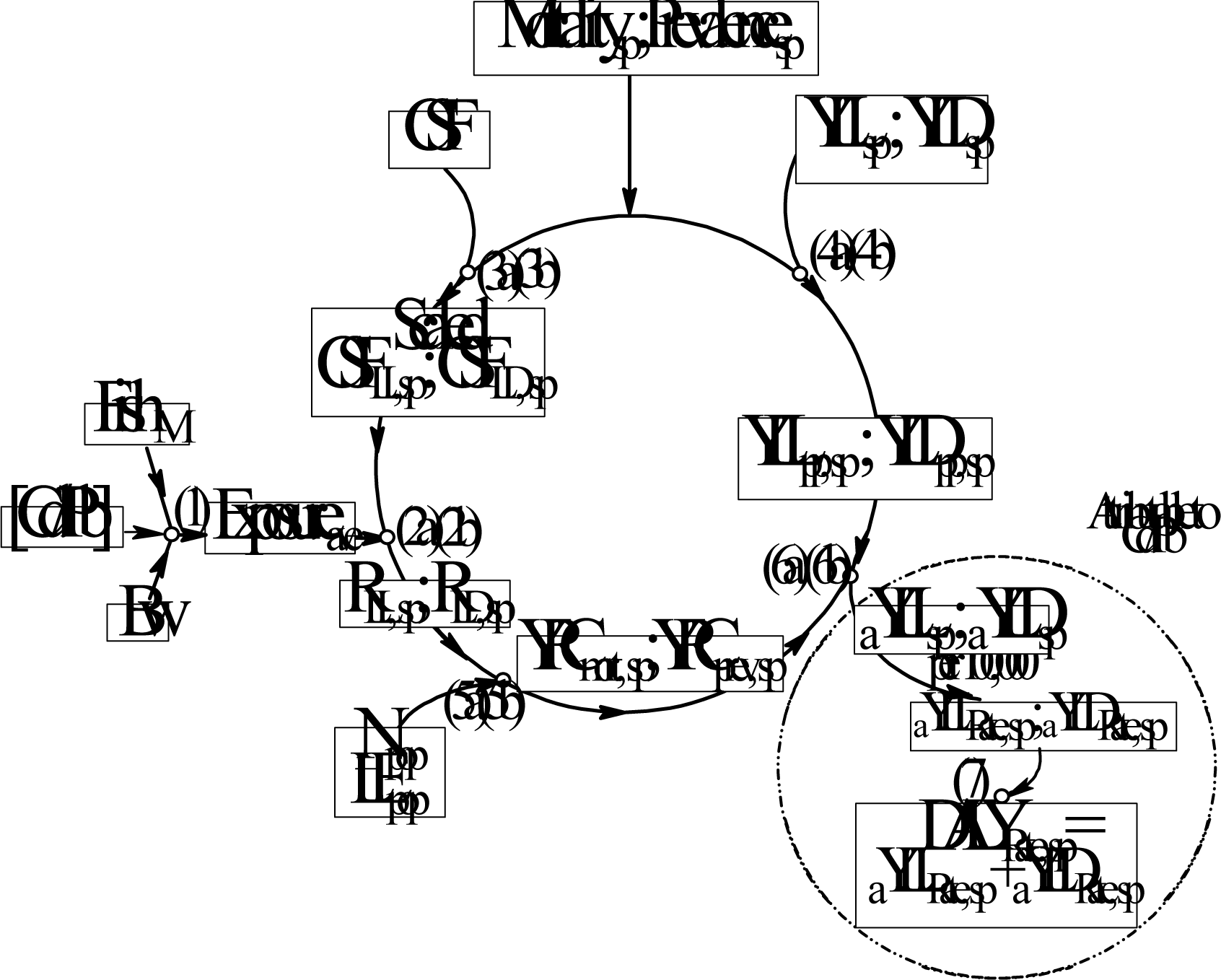
Conceptual Framework for Assessing the Health Impact of the Toxic Heavy Metals

**Evaluation of DALYs:** Equations 1 to 7 determined the attributable burdens of cancers to cadmium and lead from fish consumption. These burdens were expressed as the rate of attributable disability-adjusted life years (_a_DALY). The average exposure (E_ave_) was calculated using Equation 1, following the formula recommended by the US EPA (2015). This formula incorporates the meta-analyzed concentrations (mg/kg) of the toxic heavy metal (THMc), the mass (M_F_) of fish consumed (kg/day) and the body weight (B_W_).

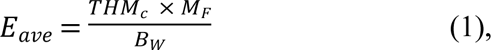

Following the calculation of average exposure, the excess lifetime cancer risks related to years of lost (R_LL_) and years lived with disability (R_LD_) were determined using Equations 2a and 2b.

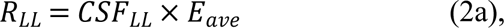

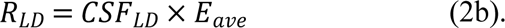

The scaled cancer slope factors for years of life lost (CSF_LL_) and years of life lived with disability (CSF_LD_) were previously calculated using Equations 3a and 3b. These factors represent the estimated increase in cancer risk per exposure unit for specific cancer endpoints. The U.S. Environmental Protection Agency (EPA) has indicated a cadmium cancer slope factor of 0.51 per mg/kg-day and a lead cancer slope factor of 0.0083 per mg/kg-day. These values represent the estimated increase in cancer risk per unit of exposure to cadmium and lead, respectively. Because cancer slope factors (CSF) vary across different cancer endpoints, specific cancer-scaled slope factors (CSF_sp_) were derived for each endpoint using Equations 4a and 4b. These equations incorporate mortality and prevalence data from 2015 to 2019.

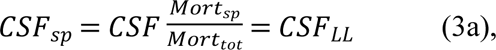

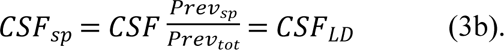

Mort_sp_ and Mort_tot_ represent specific and total mortality, while Prev_sp_ and Prev_tot_ represent the specific and total prevalence of cadmium- and lead-related cancers.

Equations 4a and 4b were used to derive the years of life lost per person (YLL_pp_) and years lived with disability per person (YLD_pp_). In these equations, YLL_sp_ and YLD_sp_ represent the specific cancer years of life lost and years of life lived with disability, respectively.

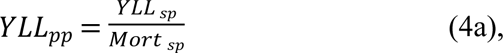

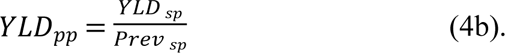

Equations 5a and 5b were used to calculate the five-year range of the specifically attributable cancers for years of life lost (Five-YRC_sp LL_) and years lived with disability (Five-YRC_sp LD_), respectively.

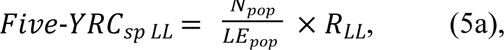

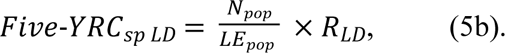

The term N_pop_/LE_pop_ represents the consumer population numbers (N_pop_) of the study area per the age-related life expectancy of that consumer population (LE_pop_), which was derived from the World Bank repository (27).

Finally, Equations 6a and 6b were used to calculate the cadmium/lead-fish-contaminated attributable to years of life lost (_a_YLL) and years of life lived with disability (_a_YLD) for each specific cancer. The corresponding rates (per 100,000 persons) were then computed as the sum of these rates, producing the specific disability-adjusted life years (DALYsp) rate, as shown in Equation 7.

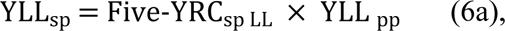

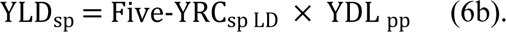

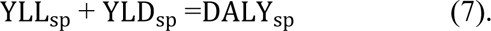

Best-fit distributions based on Chi square statistic, percentile–percentile and quantile–quantile plots were determined for each parameter using @Risk version 8.2.2010.0 (Palisade Corporation, Ithaca, NY, USA). Monte Carlo simulations were then run on the output of each equation at 100,000 iterations with @Risk version 8.2.2010.0 (Palisade Corporation, Ithaca, NY, USA).

## 3.0 Results

### 3.1 Occurrence of cadmium and lead in the fish products

Over the five-year period, cadmium was found in all samples tested, while the occurrence of lead ranged from 85% to 100%, with the consecutive years 2017 – 2019 recording 100% occurrence (Fig. 3).

**Figure 3.**
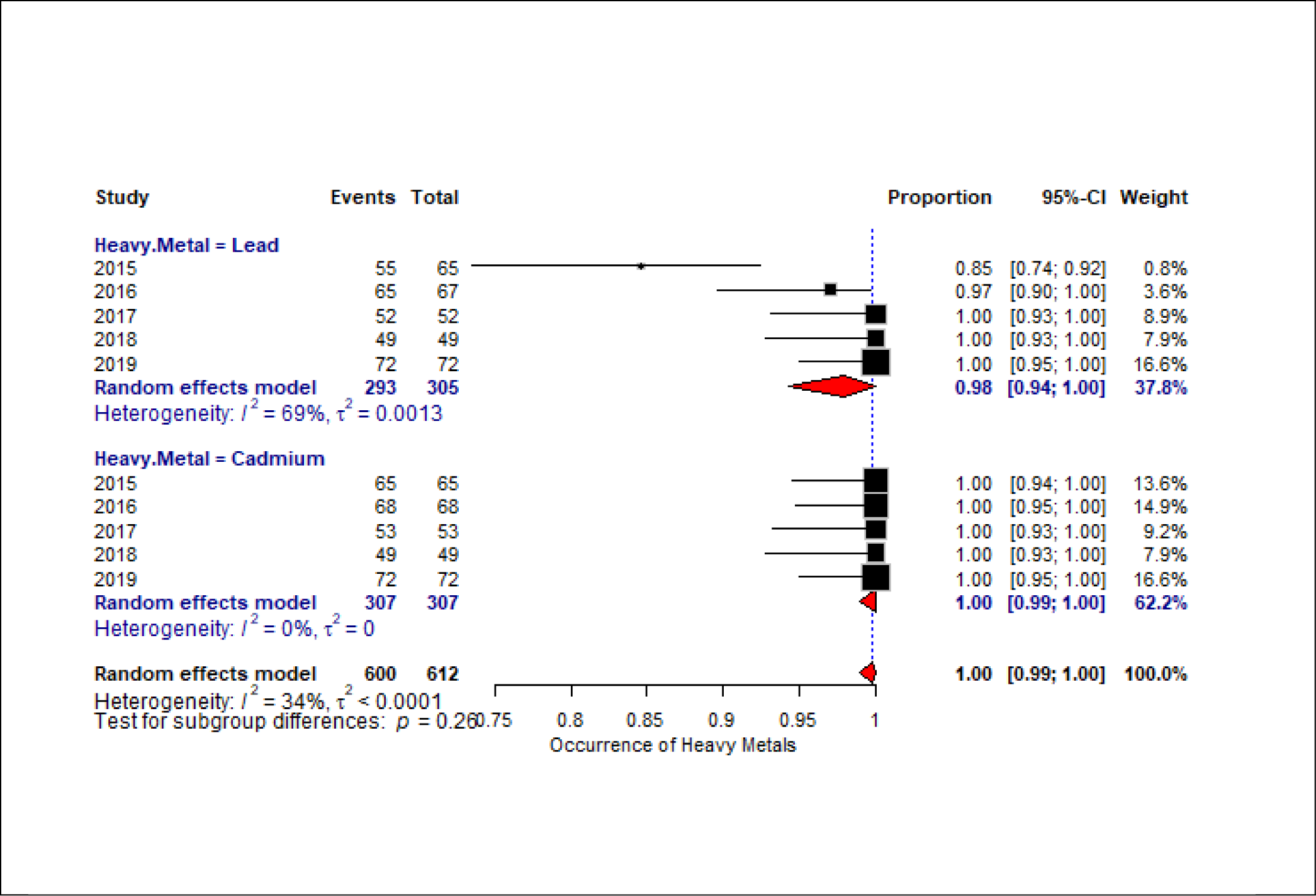
A Forest plot of the proportion of heavy metal (cadmium and lead) in tuna samples from 2015 to 2019. The number of positive samples is indicated by ‘Events’. The ratios of positive samples are indicated in the proportion column. Heterogeneity parameters are indicated by *I^2^ and τ^2^*

### 3.2 Meta-analyses of cadmium and lead concentrations

The meta-analysis of 10 studies on cadmium and lead contamination in tuna samples revealed a high prevalence of both heavy metals, with pooled proportions approaching 1.00 for both metals (Figure 3). While the overall heterogeneity among the datasets was moderate (*I^2^* =34%), subgroup differences identified no statistically significant variability (p = 0.47) between Lead and Cadmium across the study period. The observation highlights the widespread presence of these contaminants in tuna and underscores the need for further investigation and potential regulatory measures to protect public health.

### 3.3 Probabilistic concentrations and exposures

**Table 1** shows the distribution of cadmium and lead exposures in fish that landed in Ghana within the historical data collected. Median lead concentrations of 6.85×10^-2^ mg/kg (RiskTriang (- 0.02573, 0.1, 0.19108)) were higher than the median cadmium concentrations 1.10×10^-2^ mg/kg (RiskPareto (7.108, 0.01) in the samples analyzed.

**Table 1.** Probabilistic outcomes of cadmium and lead ingestions and their estimated exposures in Ghana.

| Statistics | Cadmium |  |  | Lead |  |  |
| --- | --- | --- | --- | --- | --- | --- |
|  | Concentration (mg/kg) | Mass consumed (kg/day) | Exposure (mg/kg(bw)-day) | Concentration (mg/kg) | Mass consumed (kg/day) | Exposure (mg/kg(bw)-day) |
| **Mode** | $1.00 \times 10^{-2}$ | $6.85 \times 10^{-2}$ | $1.14 \times 10^{-5}$ | $9.95 \times 10^{-2}$ | $6.85 \times 10^{-2}$ | $1.14 \times 10^{-4}$ |
| **Median** | $1.10 \times 10^{-2}$ | $6.85 \times 10^{-2}$ | $1.26 \times 10^{-5}$ | $9.10 \times 10^{-2}$ | $6.85 \times 10^{-2}$ | $1.04 \times 10^{-4}$ |

Table 1 suggests a higher lead contamination in the products, with corresponding higher exposures. Other studies have also reported generally higher concentrations of lead than cadmium in tuna (28–31).

### 3.4 Probabilistic risks of years of life lost and years of life lived with disability

Table 2a presents the risks of years of life lost (R_LL_) and years of life lived with disability (R_LD_) for prostate, bladder, kidney, lung and pancreatic cancers attributable to cadmium-contaminated fish consumed. For prostate cancer, the data presented revealed a relatively benign picture. Even among younger males aged 15-19, cadmium exposure, a known risk factor, had a negligible impact on premature mortality (0.0).

**Table 2a.** Probabilistic Modal and median Risks of Years of Life Lost and Years of Life lived in disability of prostate, kidney, pancreatic, lung and bladder cancers of some age groups in Ghana.

| Risk of Years of Life Lost from cadmium exposure |  |  |  |  |  |  |  |  |  |  |
| --- | --- | --- | --- | --- | --- | --- | --- | --- | --- | --- |
|  |  | Prostate<br>Cancer | Kidney<br>Cancer |  | Pancreatic<br>Cancer |  | Lung<br>Cancer |  | Bladder<br>Cancer |  |
| Age Group<br>(years) | Gender<br>Statistics | Male | Male | Female | Male | Female | Male | Female | Male | Female |
| 15-19 | Mode | 0.00 | $1.99 \times 10^{-9}$ | $1.47 \times 10^{-9}$ | $9.58 \times 10^{-10}$ | $1.00 \times 10^{-9}$ | $9.58 \times 10^{-10}$ | $8.41 \times 10^{-9}$ | $4.24 \times 10^{-10}$ | $3.22 \times 10^{-10}$ |
| | Median | 0.00 | $5.47 \times 10^{-6}$ | $3.98 \times 10^{-6}$ | $2.59 \times 10^{-6}$ | $2.71 \times 10^{-6}$ | $2.60 \times 10^{-6}$ | $2.27 \times 10^{-5}$ | $1.14 \times 10^{-6}$ | $8.68 \times 10^{-7}$ |
| 20-54 | Mode | $1.32 \times 10^{-7}$ | $3.94 \times 10^{-8}$ | $2.86 \times 10^{-8}$ | $1.52 \times 10^{-7}$ | $2.14 \times 10^{-7}$ | $2.51 \times 10^{-7}$ | $1.60 \times 10^{-7}$ | $1.39 \times 10^{-7}$ | $2.18 \times 10^{-8}$ |
| | Median | $3.58 \times 10^{-4}$ | $1.07 \times 10^{-4}$ | $7.73 \times 10^{-5}$ | $4.17 \times 10^{-4}$ | $5.79 \times 10^{-4}$ | $6.81 \times 10^{-4}$ | $4.30 \times 10^{-4}$ | $3.75 \times 10^{-4}$ | $5.87 \times 10^{-5}$ |
| 55-89 | Mode | $2.69 \times 10^{-6}$ | $9.45 \times 10^{-8}$ | $4.71 \times 10^{-8}$ | $5.66 \times 10^{-7}$ | $9.76 \times 10^{-7}$ | $1.02 \times 10^{-6}$ | $4.06 \times 10^{-7}$ | $2.57 \times 10^{-7}$ | $1.33 \times 10^{-7}$ |
| | Median | $7.26 \times 10^{-3}$ | $2.58 \times 10^{-4}$ | $1.27 \times 10^{-4}$ | $1.54 \times 10^{-3}$ | $2.64 \times 10^{-3}$ | $2.77 \times 10^{-3}$ | $1.10 \times 10^{-3}$ | $6.94 \times 10^{-4}$ | $3.58 \times 10^{-4}$ |
| Risk of Years of Life in Disability from cadmium exposure |  |  |  |  |  |  |  |  |  |  |
| 15-19 | Mode | 0.00 | $1.07 \times 10^{-7}$ | $3.65 \times 10^{-8}$ | $2.89 \times 10^{-9}$ | $3.16 \times 10^{-9}$ | $3.42 \times 10^{-9}$ | $8.68 \times 10^{-8}$ | $1.25 \times 10^{-8}$ | $8.26 \times 10^{-9}$ |
| | Median | 0.00 | $2.59 \times 10^{-5}$ | $2.50 \times 10^{-5}$ | $1.98 \times 10^{-6}$ | $2.16 \times 10^{-6}$ | $2.32 \times 10^{-6}$ | $2.10 \times 10^{-5}$ | $8.59 \times 10^{-6}$ | $5.70 \times 10^{-6}$ |
| 20-54 | Mode | $5.44 \times 10^{-6}$ | $9.40 \times 10^{-7}$ | $3.11 \times 10^{-7}$ | $1.58 \times 10^{-7}$ | $5.87 \times 10^{-7}$ | $3.36 \times 10^{-7}$ | $2.28 \times 10^{-7}$ | $1.85 \times 10^{-6}$ | $2.37 \times 10^{-7}$ |
| | Median | $1.32 \times 10^{-3}$ | $2.29 \times 10^{-4}$ | $2.16 \times 10^{-4}$ | $1.09 \times 10^{-4}$ | $1.43 \times 10^{-4}$ | $2.29 \times 10^{-4}$ | $1.57 \times 10^{-4}$ | $1.28 \times 10^{-3}$ | $1.63 \times 10^{-4}$ |
| 55-89 | Mode | $3.89 \times 10^{-5}$ | $2.35 \times 10^{-7}$ | $4.01 \times 10^{-7}$ | $1.05 \times 10^{-6}$ | $6.54 \times 10^{-7}$ | $2.50 \times 10^{-6}$ | $3.37 \times 10^{-7}$ | $8.87 \times 10^{-7}$ | $3.90 \times 10^{-7}$ |
| | Median | $9.39 \times 10^{-3}$ | $1.61 \times 10^{-4}$ | $9.77 \times 10^{-5}$ | $2.55 \times 10^{-4}$ | $4.54 \times 10^{-4}$ | $6.00 \times 10^{-4}$ | $2.34 \times 10^{-4}$ | $6.12 \times 10^{-4}$ | $2.71 \times 10^{-4}$ |

For males aged 20-54, the R_LL_ and R_LD_ of bladder cancer (attributable to cadmium,) were minimal (10^-10^ -1.28×10^-3^), suggesting that while a severe health concern, bladder cancer does not pose a significant risk for the specific demographic group. However, cadmium-contaminated fish attributable to prostate cancer indicated a stark contrast to bladder cancer, emerging as a more formidable health challenge with substantially higher R_LL_ and R_LD_ values (10^-7^-7.26×10^-3^) across all the age groups between 20 and 89. The observation of a greater risk of attributable cadmium/lead-fish contamination for both premature death and disability from cancers is a concerning finding. This observation indicates the importance of targeted prevention and early detection strategies for prostate cancer in Ghana. Pancreatic cancer exhibited higher risks (10^-10^- 2.64×10^-3^) than bladder cancer (10^-10^-1.28×10^- 3^) but lower risks than prostate cancer (10^-7^- 7.26×10^-3^). While pancreatic cancer remains a serious concern, the data suggest that it may be less of a priority compared to prostate cancer in terms of public health interventions. However, the fact of the risk for cancer from this food-hazard pair is of greater importance than the differences in risk levels of the type of cancer. Therefore, the corresponding risk management efforts should be targeted at minimizing exposure to the toxic metals in tuna, rather than reducing particular types of cancer.

In Table 2b, we observe that though lead exposure, from literature, has a potential health risk, the low median risk values (10^-11^-10^-8^) of kidney cancer in this current study do not pose any significant threat in the younger age group. Generally, the R_LL_ due to kidney cancer increases from middle to older age groups, with values approximately rising from 10^-9^ to 10^-2^. The R_LD_ is higher for the middle age group (10^-6^) than the elderly (10^-7^). However, the consistently low median R_LL_ and R_LD_ risk values for both cancers suggested a limited impact on both premature mortality and disability challenges. It is worth noting that among the cancers analyzed in both Tables 2a and 2b, lead contamination in fish in attributable stomach cancer in Ghanaian males aged 55-89 exhibited the highest median R_LL_ (1.43×10^-2^).

**Table 2b.** Probabilistic Modal and median Risks of Years of Life Lost and Years of Life lived in disability of kidney and stomach cancers of some age groups in Ghana.

| Risk of Years of Life Lost from lead exposure |  |  |  |  |  | Risk of Years of Life in Disability from lead exposure |  |  |  |
| --- | --- | --- | --- | --- | --- | --- | --- | --- | --- |
| Age Group (years) | Gender Statistics | Kidney cancer |  | Stomach cancer |  | Kidney cancer |  | Stomach cancer |  |
|  |  | Male | Female | Male | Female | Male | Female | Male | Female |
| 15-19 | Mode | $7.32 \times 10^{-11}$ | $5.34 \times 10^{-11}$ | $7.18 \times 10^{-8}$ | $7.70 \times 10^{-11}$ | $5.97 \times 10^{-8}$ | $5.24 \times 10^{-8}$ | $1.29 \times 10^{-8}$ | $1.56 \times 10^{-8}$ |
| | Median | $6.71 \times 10^{-8}$ | $4.88 \times 10^{-8}$ | $6.45 \times 10^{-5}$ | $7.11 \times 10^{-8}$ | $1.55 \times 10^{-7}$ | $1.49 \times 10^{-7}$ | $3.64 \times 10^{-8}$ | $3.45 \times 10^{-8}$ |
| 20-54 | Mode | $1.44 \times 10^{-9}$ | $1.04 \times 10^{-9}$ | $4.71 \times 10^{-6}$ | $8.08 \times 10^{-9}$ | $5.48 \times 10^{-7}$ | $4.68 \times 10^{-7}$ | $3.89 \times 10^{-7}$ | $5.74 \times 10^{-7}$ |
| | Median | $1.32 \times 10^{-6}$ | $9.51 \times 10^{-7}$ | $4.23 \times 10^{-3}$ | $7.50 \times 10^{-6}$ | $1.36 \times 10^{-6}$ | $1.29 \times 10^{-6}$ | $1.01 \times 10^{-6}$ | $1.55 \times 10^{-6}$ |
| 55-89 | Mode | $3.48 \times 10^{-9}$ | $1.72 \times 10^{-9}$ | $1.59 \times 10^{-5}$ | $2.71 \times 10^{-8}$ | $3.97 \times 10^{-7}$ | $2.61 \times 10^{-7}$ | $7.17 \times 10^{-7}$ | $1.24 \times 10^{-6}$ |
| | Median | $3.18 \times 10^{-6}$ | $1.58 \times 10^{-6}$ | $1.43 \times 10^{-2}$ | $2.51 \times 10^{-5}$ | $9.57 \times 10^{-7}$ | $5.81 \times 10^{-7}$ | $2.03 \times 10^{-6}$ | $2.97 \times 10^{-6}$ |

### 3.5 The five-year cancer prevalence rates and the elements of the burden of cancers

Table 3a presents the five-year range of cancer (Five-YRC) prevalence rates attributable to cadmium-contaminated fish in consumers’ diets. Analysis across different age groups and genders revealed that, for gender-specific patterns, prostate cancer was the most prevalent type among males, presenting median values ranging between 748.92 in the middle-aged group to 5,341.90 in the aged group. These represented the highest prevalences across all age groups. Bladder cancer also poses a significant threat to male health, presenting median values ranging between 4.91 to 346.66. In contrast, females presented significant Five-YRC prevalent values (1.13-236.30) relating to the pancreas, which consistently ranked as the most prevalent cancer among females in the study area.

**Table 3a:**
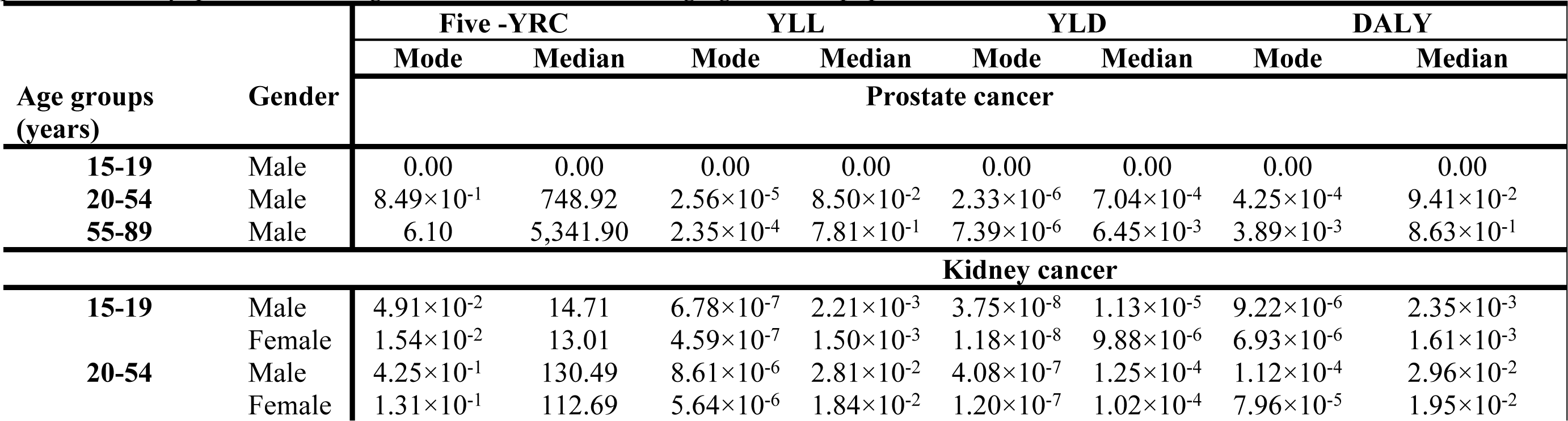
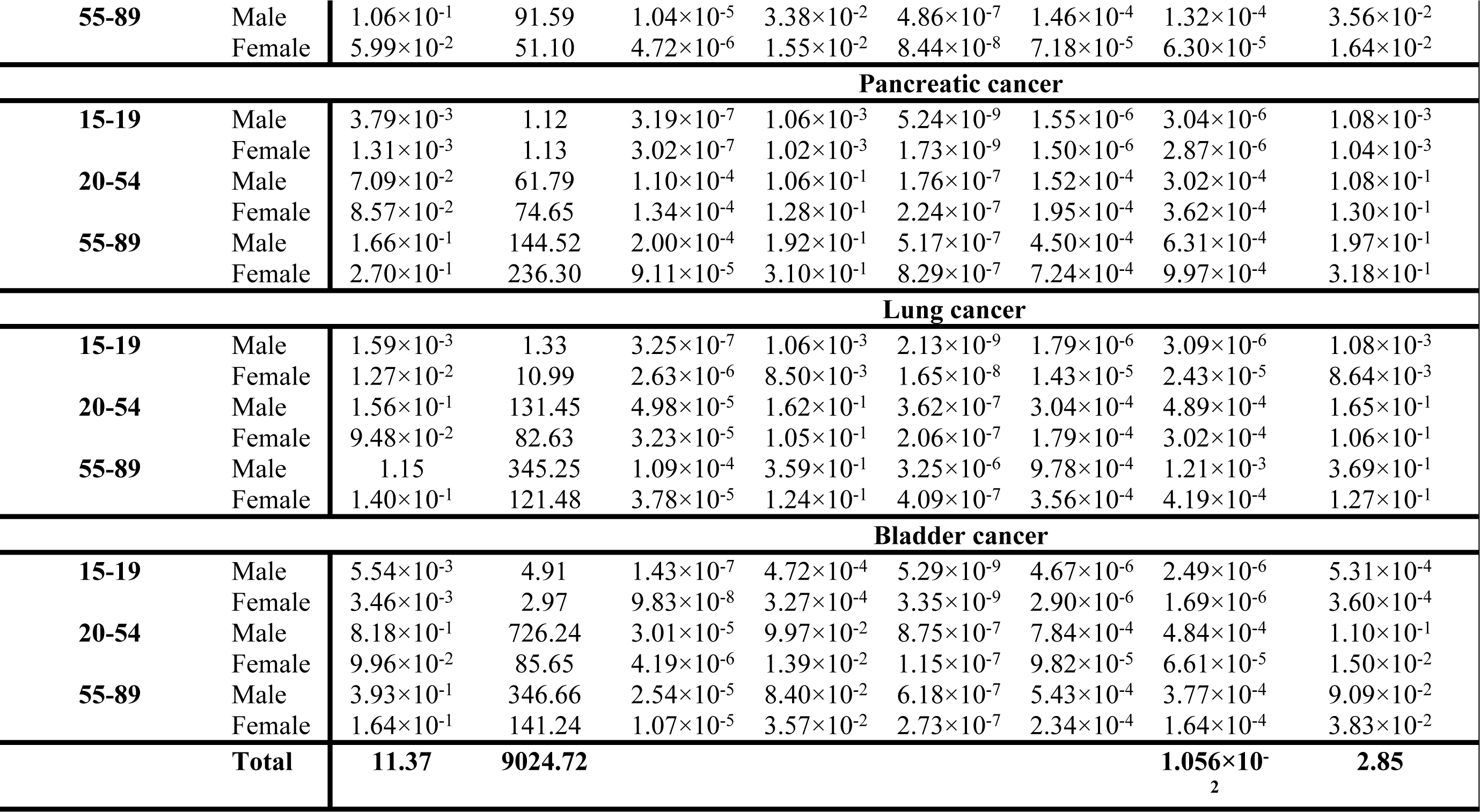
The probabilistic Cadmium-induced five-year cancer prevalence rates and the elements of the burden of cancers in prostate, kidney, pancreatic, lung and bladder cancer among age-related populations in Ghana.

Age-related trends were evident across all cancer types studied. The middle-aged group (20-54years) presented a significantly higher Five-YRC prevalence for both kidney cancer (females: 112.69, males: 130.49) and bladder cancer (males: 726.24) compared to the older adults (55-89years) age group. Thus, this middle-aged group is more likely to develop cadmium-attributable cancers than younger and older individuals. Generally, the older age group presented significant Five-YRC prevalences for prostate cancer in males, pancreatic and lung cancers in both males and females and bladder cancer in females.

A subtle observation was noted: the Five-YRC prevalences of pancreatic cancer (236.30) among older females (55-89 years) was significantly higher (144.52) than that of their male counterparts. This observation suggests that exposure patterns or risk factors may disproportionately affect women in this age group.

The DALYs revealed significant disparities across age groups. While the DALY was relatively low (10^-6^-10^-3^) among younger individuals (15-19 years), it increased substantially (10^-5^-10^-4^) in the 20-54 age group, reflecting the impact of early-onset disease. The highest DALY values (10-^5^-10^-3^) were observed in the 55-89 age group, underscoring the substantial burden of cancer among older adults. Distinct patterns emerged regarding the contributions of YLL and YLD to the overall DALY. In younger age groups, the YLL frequently played a more significant role, ranging from 10^-8^ for bladder cancer to 10^-1^ for prostate, pancreatic and lung cancer.

An analysis of the Five-YRC prevalence rates in Table 3b revealed several important trends for exposure to lead-contaminated fish products across different age groups and genders. For the age group 15-19, stomach cancer consistently exhibited a lower (10^-3^-10^-2^) Five-YRC prevalence rate than kidney cancer (10^-2^), regardless of gender. In the 20-54 age group, genders played a significant discriminatory role in the Five-YRC prevalence of lead-contaminated fish-attributable cancers. Kidney cancer exhibited the highest rate, particularly among males, while stomach cancer was more prevalent among females. Males with kidney cancer had a rate of 0.76, exceeding that of females (0.66). Conversely, females with stomach cancer had a rate of 0.79, surpassing the male rate of 0.56.

**Table 3b:**
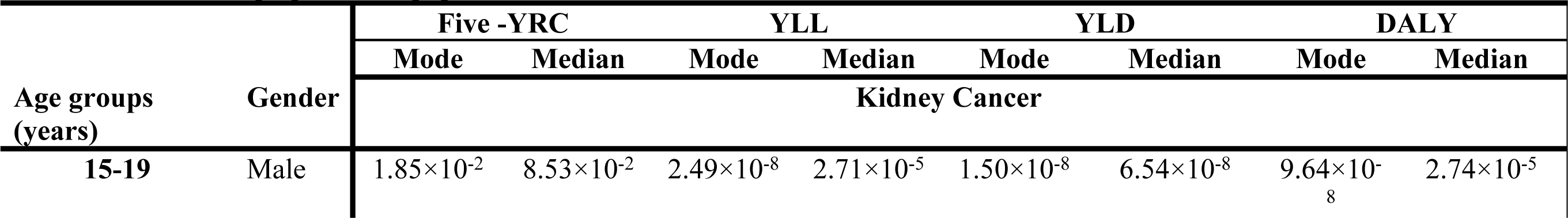
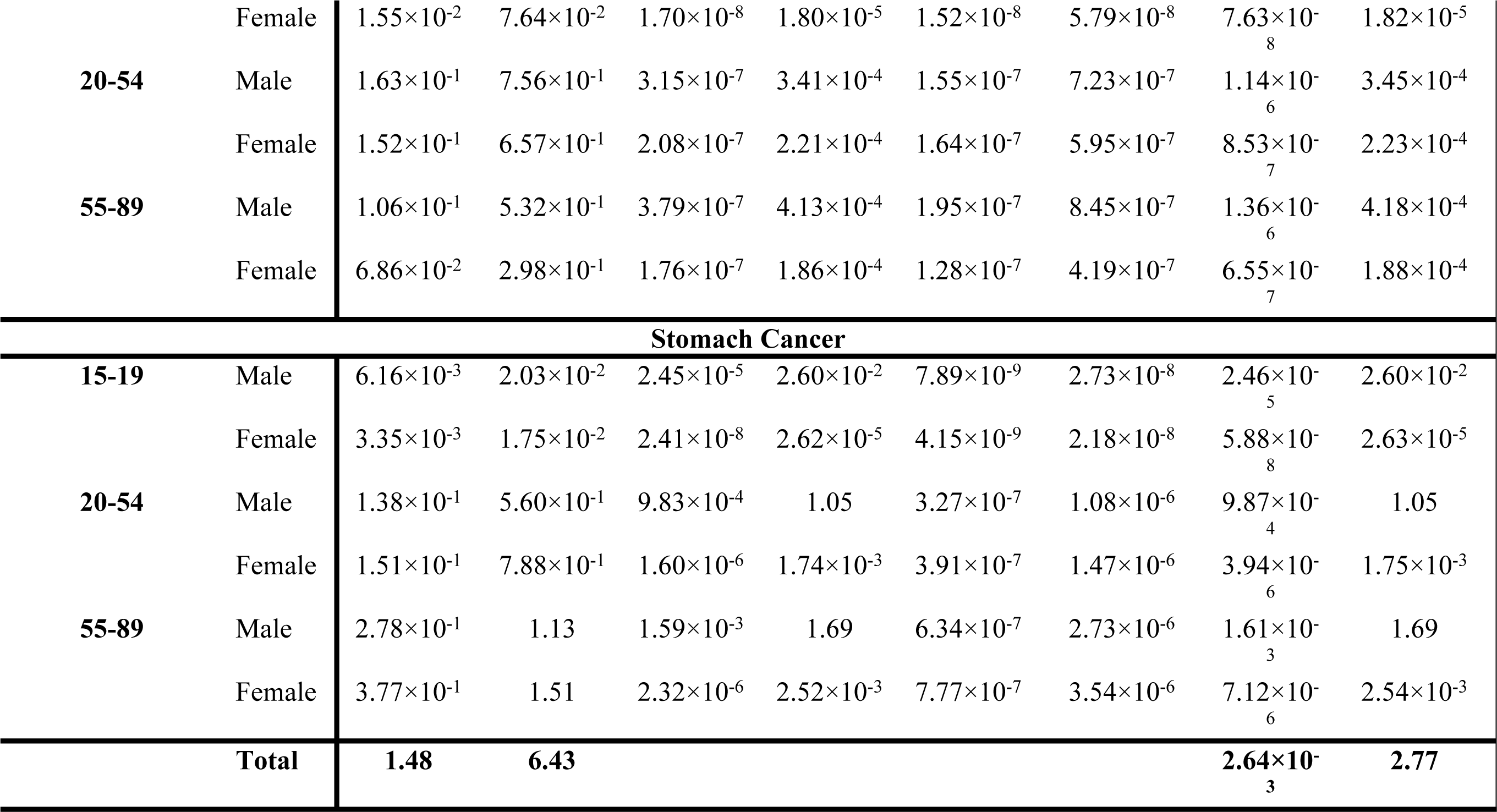
The probabilistic Lead-induced five-year cancer prevalence rates and the elements of the burden of cancers in kidney and stomach cancers among age-related populations in Ghana.

In the 55-89 age group, stomach cancer was the most prevalent cancer among both genders. Aside from kidney cancer, there is an apparent age-related increase in the Five-YRC prevalence of stomach cancer, rising from 3.35×10^-3^ in the youngest age group (15-19) to 0.79 in the middle age group (20-54years) and then to 1.51 in the oldest age group (55-89years). The middle age group presented higher Five-YRC rates than the older age group. Gender differences in the prevalence of these cancers were also significant. Females presented higher values (1.51) compared to their male counterparts (1.13), indicating that both males and females were significantly affected by the risk of developing stomach cancer.

For the 15-19 age group, the DALY was relatively low (10^-8^-10^-2^) compared to the other age groups, primarily due to lower Five-YRC prevalence rates (10^-3^-10^-2^) among this younger population. DALY values were affected by both age-related factors and gender. For the 20-54 age group, both stomach and kidney cancer presented higher rates than in the younger age group, reflecting the growing prevalence of Lead-attributable cancer along the ageing curve. However, males exhibited significantly higher DALY values (1.05) than females (1.75×10^-3^). The highest DALY values were observed in the 55-89 age group for stomach cancer in both genders, emphasizing the substantial burden of cancer among older individuals. As with cadmium-contaminated fish products, YLL contributed more significantly to the DALY than YLD in all age groups.

## 4.0 Discussion

The study sought to determine the levels of cadmium and lead in selected tuna samples that landed in Ghana, assess the potential health risks associated with consuming this contaminated tuna, and inform public health policy and consumer awareness in Ghana regarding the dangers of heavy metal exposure through tuna consumption. The study revealed that lead concentrations in fish landed in Ghana were significantly higher than cadmium concentrations. Specifically, the lead concentrations had a mode of 9.95×10^-2^ mg/kg, while the cadmium concentrations had a mode of approximately 1.00×10^-2^ mg/kg. This disparity was reflected in the daily exposure levels.

Concentrations of metallic contaminants as indicated in Table 1, shows that lead concentration 9.10×10^-2^ was higher than cadmium 1.10×10^-2.^ This study’s findings align with other studies. For instance, (32) reported the presence of heavy metals in canned fish products sold in Ghana, with some samples exceeding the recommended EU safety limits for lead (0.30 mg/kg and cadmium 0.10 mg/kg) (33). Also, another study in the eastern Pacific region analyzed 2,572 tuna samples between 2009 and 2016 (34). Though the mean metal concentrations were reported to be low (0.03 mg/kg for cadmium and 0.05 mg/kg for lead), random samples from local stores showed even lower concentrations. Over 0.58% of samples had readings near or above the limits, often due to cross-contamination. However, the study concluded that Ecuadorian canned tuna is safe for consumption. Lead exposure was considerably greater than cadmium exposure, presenting median values of 1.04×10^-4^ mg/kg(bw)-day, compared to 1.26×10^-5^ mg/kg(bw)-day. Although both contaminants were present in the fish, the higher levels of lead and the associated exposure posed a greater health risk. In a study by (35), it was mentioned that the synergistic effect or in some cases, these antagonistic effects of heavy metal accumulation in fish must be explored to make comprehensive decisions on fish contamination.

The findings of this study are particularly concerning, given the potential health risks associated with chronic heavy metal exposure. Chronic exposure to high levels of these contaminants can lead to various health problems, including neurological disorders, kidney damage, and cancer (36). The current study and other studies’ findings underscore the importance of continuous monitoring and regulation to ensure consumer safety and minimize health risks associated with heavy metal ingestion from fish products. This approach includes quality and stricter regulations on fishing practices, improving water quality, and promoting sustainable fishing methods. The Ecuadorian study on canned fish gives credence to the call for public awareness campaigns to educate consumers about the potential health risks associated with heavy metal exposure and guide safe fish consumption.

### 4.1 Cadmium and lead in landed tuna and public health risks

From Table 2a, the study indicated that cadmium exposure from landed tuna consumption is associated with higher YLL and YLD from prostate cancer compared to bladder or pancreatic cancer, particularly among younger males. However, while pancreatic cancer poses a significant risk, prostate cancer appears to be a more pressing public health concern among the consumer population in Ghana. The strong association between cadmium and prostate cancer is a global health concern, and this study further underscores the adverse effects of cadmium exposure. (37) previously highlighted the various pathways to cadmium exposure, including occupational settings, smoking, and dietary intake. These findings have been corroborated by a population-based cohort study of 41 089 Swedish men aged 45– 79 years from 1998 through 2009. The study findings provided support that dietary cadmium exposure may have a role in prostate cancer development (38).

Conversely to our findings, A Danish cohort study involving 26,778 men found no significant association between dietary cadmium intake and prostate cancer risk, irrespective of disease aggressiveness or modifying factors like smoking and zinc intake (39)

In Table 2b, we indicate that lead exposure from fish consumption is associated with low risks of YLL and YLD from kidney cancer, with the highest risk observed among elderly males aged 55-89. However, the overall risk of lead exposure on both YLL and YLD challenges is limited. Our findings regarding lead exposure from fish consumption align with a recent Central Asian study (40). Their study examined mercury, cadmium, and lead levels in fish and seafood and found that cadmium and lead were generally detected in lower concentrations than Hg. While exposure to all three metals is a concern, the authors concluded that mercury poses the most significant risk to human health in the region.

Contrary to the assumption that elderly adults are usually at risk, recent research suggests that elderly adults may not always be the primary population at risk of cadmium exposure. A study conducted in Zhejiang, China, from 2018 to 2022 analyzed cadmium levels in aquatic products and found that marine crustaceans were the primary source of exposure. While the average cadmium exposure for residents was below the provisional tolerable monthly intake, young children aged 2-3 had significantly higher exposures (41). This observation highlights the need for targeted interventions to address cadmium exposure risks in specific age groups and dietary habits.

### 4.2 Cancer burden

Our study found a link between cadmium exposure from fish consumption and high five-year cancer prevalence rates, particularly in middle-aged individuals. Prostate cancer (0.85), kidney cancer (0.13-0.43), pancreatic cancer (0.07-0.086), lung cancer (0.095-0.160), and bladder cancer (0.10-0.80) were all associated with increased cadmium exposure. Among these cancers, prostate cancer was most prevalent in men (0.85), while kidney cancer was more common in women (0.13). Our findings are consistent with a systematic review and meta-analysis of cohort studies examining the relationship between fish consumption and prostate cancer risk or mortality (42). Their study, which collected reports from Sweden, the U.S., the U.K., and Japan, revealed a correlation between consuming various types of fish and an elevated risk of prostate cancer. In their paper, a total of 44,722 prostate cancer prevalences were reported, including 5,422 advanced cases, 3,779 deaths, 613 cases of progression, and 406 localized cases. In comparison to our study, our reported numbers are significantly lower.

Our study has indicated that the DALYs associated with cadmium exposure from fish consumption increase with age. The findings support a review study that shows foodborne diseases, including heavy metal-related such as cadmium, significantly contribute to the global disease burden, especially in low-and middle-income countries (43). The review estimated that studies on four heavy metals, including cadmium, found over 9 million DALYs lost in six countries. Our study found the highest DALYs in the oldest age group (55-89), suggesting a significant cancer burden among older adults. This observation aligns with the review’s finding that older populations bear a disproportionate burden. In younger age groups, our study found premature death (YLL) contributing more significantly to overall DALY. This finding supports the notion that younger age groups may experience more premature deaths.

Again, the present study found that stomach cancer arising from heavy metal exposure in tuna is the most prevalent cancer among both genders in the older age group (55-89). Indeed, gastric cancer is the 5th most common and the third deadliest cancer globally (44). The 5-year survival rate is 31% in the U.S. when diagnosed at the regional stage, which has better outcomes than at the last stages (45). For these reasons, cancer from cadmium-contaminated fish remains a public health concern. Our study shows that the Five-YRC prevalence of stomach cancer increases with age, and women have higher rates than men, unlike advanced countries where gastric cancer is typically more prevalent among males (46). It has been suggested that low-and middle-income consumers are more likely to develop gastric cancer. Also, lower socioeconomic status is often linked to higher *H. pylori* risk, which is critical in the development of stomach cancer (47).

Concerning lead contaminated fish, the current study found that DALYs increased with age, highlighting that age can influence the disease burden. This information aligns with another study emphasizing the impact of DALY-related heavy metal exposure (48).

### 4.3 Public health

Our findings suggest that cadmium and lead contamination in tuna translate to varying degrees of prostate, bladder, stomach, pancreatic, and kidney cancer, with disparities in risk levels for genders and age groups (Tables 2a, 2b, 3a, and 3b). Other studies corroborate that cancer risks from dietary cadmium and lead are unevenly distributed, influenced by factors such as exposure route, cancer subtype, and hormonal interactions, requiring tailored public health strategies (49–53).

Dietary interventions can be crucial in mitigating the risks associated with cadmium and lead exposure.

### 4.4 Limitations

The study relied on a small sample size, which might pose a significant limitation, as it may not accurately represent the broader population. While a meta-analysis assessed how robust or biased the data used for the study may be, the associated uncertainty in statistical power could limit definitive conclusions about the outcomes of the health impact of cadmium and lead health exposures. Also, using varying data sources could introduce inconsistencies resulting from the original collection methods and conditions. This inconsistency in historical contaminant concentration data can complicate analysis and interpretation.

Methodological challenges, such as the lack of control groups, may have complicated the study, however, a non-controlled design can be feasible for observing long-term effects. While it may be challenging to isolate the specific effects of the intervention due to the potential influence of confounding variables, long-term studies can reveal trends and patterns that may not be apparent in shorter-term studies. Thus, with the robust data analysis techniques employed in this study, valuable outcomes can still be gained. It is also possible that geographical, seasonal, and processing-related factors could influence contaminant levels in fish, which may not be fully accounted for in the analysis. The findings may not be universally applicable, as the risks associated with cadmium and lead exposure may vary across different populations over the years.

Despite these limitations, the study offers valuable insights into the health risks associated with 2cadmium and lead exposure from specific fish. However, it is important to interpret the results cautiously and consider the limitations enumerated when drawing conclusions and formulating public health recommendations.

## 5.0 Conclusion

The findings of the study suggest that for tuna consumers in Ghana, cadmium and lead contamination may pose risks for prostate, bladder, stomach, pancreatic, and kidney cancer. However, the levels of risk vary for cancer types, gender, and age groups of consumers. Targeted risk management actions are required to minimize public exposure to the hazards. Interventions such as surveillance of heavy metal occurrence in fish landings, public and industry education on the prevention of water body contamination, safe fishing practices, enforcement of regulations on fish safety, and regimentation of the fish industry may support risk reduction.

## Data Availability

The Raw data for this manuscript is deposited in a repository here.

## Data Availability

All relevant data are within the manuscript and its Supporting Information files.

https://swpgh-my.sharepoint.com/:f:/g/personal/jessica_nkansah_gsa_gov_gh/ElgL-hTf8rVItGLsM6n4besBJY5w6iu3s7nUfXb6JusWjg?e=8utpea

## Acknowledgement

We acknowledge the contributions of all who helped in collating the retrospective data Mr. Osborn Mensah, Mr. Samuel k. Tweneboah Koduah, Mrs. Betty Giftan, and Mr. Kwabena Sakyi

